# Evaluation of non-sputum-based diagnostics for pediatric tuberculosis: the Pediatric TB Diagnostic (PDTBDx) cohort protocol

**DOI:** 10.64898/2026.04.01.26350011

**Authors:** Brendan Mullen, Joy Githua, Jaclyn N. Escudero, Jerphason Mecha, Lucy Kijaro, Maureen Ndunge, Moses Muriithi, Isaac Kibet, Grace John-Stewart, Elizabeth Maleche-Obimbo, Videlis Nduba, Sylvia M. LaCourse, the PDTBDx study team

## Abstract

Tuberculosis (TB) is a significant cause of morbidity and mortality in children and adolescents, causing 172,000 deaths in 2024 in children and adolescents worldwide. Diagnostic challenges are pronounced in pediatrics, in which collecting respiratory specimens is challenging and TB is often paucibacillary, leading to delayed diagnosis and increased mortality. We describe the protocol and methodology of the Pediatric TB Diagnostic (PDTBDx) cohort, a study with the primary aim of evaluating non-sputum-based TB diagnostics for diagnosis and treatment response in children. This is a prospective observational cohort study of >400 children recruited from inpatient and outpatient clinical sites in Nairobi, Kenya. Children <15 years presenting to study clinical sites with TB symptoms will be considered for enrollment as symptomatic participants. Enrolled participants will undergo rigorous clinical assessment and longitudinal follow-up to ensure appropriate diagnostic classification by NIH consensus statement guidelines for pediatric TB. Baseline evaluation includes symptom assessment, chest x-ray, HIV testing, respiratory TB culture and GeneXpert Ultra, and urine LAM. Subsequent visits occur at week 2, months 1, 2, 4, 6,12 and 24. Blood and urine specimens will be collected at baseline and at follow-up visits for storage for evaluation of novel diagnostic assays, including exosome-based and CRISPR-based TB biomarkers. This large, prospective cohort of pediatric participants with and without TB follows a consistent and rigorous protocol for diagnosing childhood TB, in concordance with internationally recognized guidelines. Assays evaluated in PDTBDx will guide improved diagnostic strategies for pediatric TB.

## Introduction

Tuberculosis (TB) remains a significant cause of morbidity and mortality worldwide. In 2024, there were 1.23 million deaths attributable to TB worldwide, with 172,000 deaths in children and adolescents <15 years old, with 80% of these deaths in children <5 years of age^1^. Over 96% of TB deaths in children occur in those not on treatment, largely due to missed or delayed diagnosis^2^. Sub-Saharan Africa carries a disproportionate burden of pediatric TB, accounting for roughly a quarter of the global childhood TB burden^1,3^. In Kenya in 2023, there were 12,884 children aged 0-14 years with reported TB, which represented 13.3% of all individuals diagnosed with TB^4^.

Existing methods of diagnosing TB typically rely on detection of *M. tuberculosis* in respiratory specimens through microbiologic or molecular methods. However, collection of high quality, diagnostic respiratory samples in children remains challenging. Additionally, TB in children is frequently paucibacillary and extrapulmonary, adding to the challenge of obtaining a microbiologic diagnosis^5^. Due to these challenges, diagnoses of TB in children and subsequent treatment decisions are often made on the basis of clinical criteria^6^. Thus, there is a significant need for the development and evaluation of novel methods of diagnosing TB in children^7^. Due to childhood TB disease having an imperfect gold standard, it is critical that the assessment of new diagnostics occurs in a well-characterized cohort with adequate follow-up to rule in or rule out TB disease^5,8^.

Here we describe the protocol and methodology for the Pediatric TB Diagnostic (PDTBDx) cohort, a prospective cohort of children with suspected TB enrolled from outpatient and inpatient sites in Nairobi, Kenya. This cohort leverages standardized TB symptom and exposure screening with post hoc categorization using consensus definitions, radiographic evaluation, and longitudinal collection of clinical specimens for routine and novel TB diagnostics, with the aim of evaluating the diagnostic performance of routine and novel TB diagnostic methods.

## Materials and Methods

### Study Design & Setting

PDTBDx is a prospective, longitudinal observational cohort that will enroll children ≤15 years with presumed tuberculosis (TB) and follow them for up to 24 months, with the primary aim of evaluating the diagnostic performance of novel and existing assays in children with presumptive TB (**Figure 1**). The study draws on methodological principles from the Regional Prospective Observational Research in Tuberculosis (RePORT) initiative, which promotes harmonized TB cohort protocols globally^9^.

**Figure 1.**
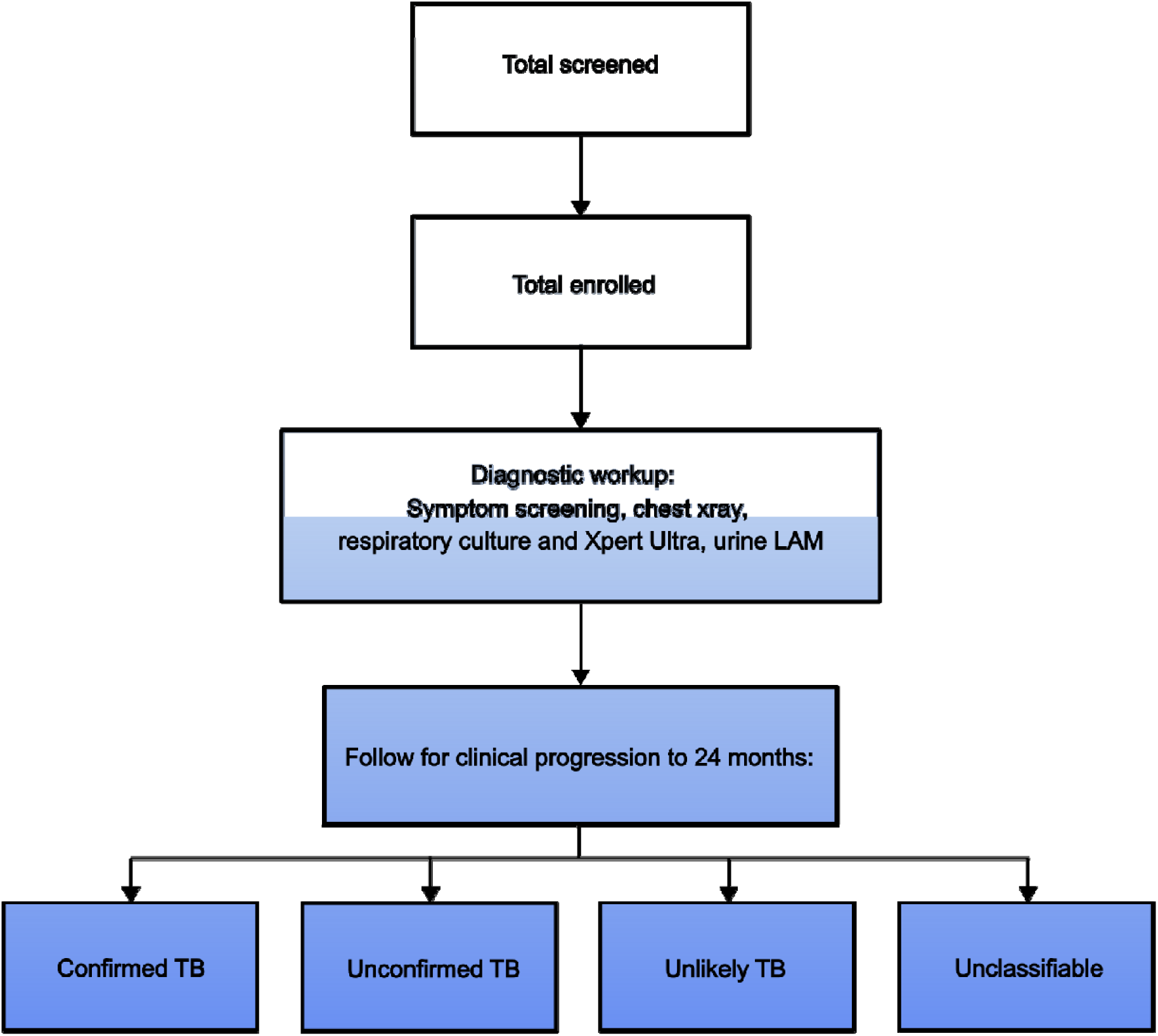
Recruitment and case classifications

Children will be recruited from inpatient wards and outpatient clinics within the National Tuberculosis Program and affiliated HIV care clinics across Nairobi, Kenya. These facilities include government TB clinics, community-based clinics, and referral sites within the county TB and HIV care network, enabling enrollment of children with a broad spectrum of TB symptoms and disease severity.

Following initial screening at these recruitment sites, eligible children will be referred to the Centre for Respiratory Diseases Research (CRDR) at the Kenya Medical Research Institute (KEMRI) for enrollment. At CRDR, participants will undergo standardized clinical assessments, detailed diagnostic evaluation, and specimen collection as outlined in the study protocol. Longitudinal follow-up at predefined intervals facilitates accurate TB disease classification and evaluation of diagnostic test performance. Participants began enrollment in May 2022. All diagnostic activities and reporting adhere to the Standards for Reporting of Diagnostic Accuracy Studies (STARD) guidelines^10^.

### Participant Eligibility

Children will be considered eligible for participation if ≤15 years of age with presumptive TB (TB symptoms) and written informed consent can be obtained from parents/guardians (with assent from children ≥13 years). Inclusion criteria include clinical signs and symptoms suggestive of TB disease (persistent cough, fever, unintended weight loss or failure to thrive, fatigue, night sweats), chest radiographic findings consistent with TB and/or microbiologic confirmation by sputum or gastric aspirate culture or Xpert Ultra, and documented HIV status or willingness to undergo HIV testing per national guidelines (**Table 1**). Children will be excluded if they have received >7 days of anti-TB treatment within 30 days prior to enrollment, plans to relocate that would interfere with 24-month study follow-up, or active conditions that would preclude informed consent or study adherence.

**Table 1.** Inclusion and Exclusion Criteria for PDTBDx Study.

| Inclusion Criteria | Exclusion Criteria |
| --- | --- |
| 1) Consent and Assent (if applicable): signed written consent/assent, or witnessed oral consent/assent in the case of illiteracy, before undertaking any study-specific activity.<br>2) Signs and Symptoms: Suspicion of TB disease (one or more criteria):<br>a. Persistent cough<br>b. Hemoptysis<br>c. Fever<br>d. Unintended weight loss or failure to thrive<br>e. Fatigue or lethargy<br>f. Night sweats<br>g. Pleuritic chest pain<br>3) Chest radiograph (CXR) findings consistent with TB, and/or sputum culture or Xpert positive.<br>4) Documentation of or willingness to be tested for HIV infection.<br>5) Agrees to the collection and storage of blood, urine, and sputum specimens for use for future research. | 1) Children of 15 years of age or more<br>2) Received >1 week (daily or intermittent doses) of any drugs with anti-TB activity within 30 days prior to provisional enrollment, including:<br>a. Any drug or combination of drugs typically used in a multidrug anti-TB therapy (isoniazid, rifampicin, pyrazinamide, ethambutol);<br>b. Any fluoroquinolone (e.g., ofloxacin, ciprofloxin, levofloxacin, moxifloxacin, nalidixic acid, sparfloxacin, and gatifloxacin);<br>c. Any other drugs with anti-TB activity (e.g., clofazamine, aminoglycosides [amikacin, kanamycin], or capreomycin).<br>3) Plans to move from his/her current residence, which would interfere with the participant's ability to complete all study visits (through the 24-Month Post-Treatment Visit). |

### Study Procedures

Participants will undergo a baseline evaluation followed by scheduled follow-up visits at week 2 and at months 1, 2, 4, 6, 12, and 24. Treatment for TB will be initiated at the treating clinician’s discretion per national guidelines, typically 6 months of treatment for drug susceptible TB or 4 months of treatment for non-severe, smear-negative disease. **Table 2** describes data and sample collection that will be obtained at baseline and follow-up visits. A subgroup of participants will be enrolled in a post-TB sub-study with additional assessments after completion of TB treatment.

**Table 2.** Schedule of Visits and Assessments.

| Activities | Visit |  |  |  |  |  |  |  |  |
| --- | --- | --- | --- | --- | --- | --- | --- | --- | --- |
|  | SCREENING | BASELINE | WEEK 2 (+/- 7 days) | MONTH 1 (weeks 3-7) | MONTH 2 (weeks 8-12) | MONTH 4 (weeks 13-17) | MONTH 6 (weeks 20-30) | MONTH 12 (weeks 40-60) and up to 24 months | MONTH 18 (-4wks and up to 24 months) |
| Informed consent | X |  |  |  |  |  |  |  |  |
| Eligibility assessment | X |  |  |  |  |  |  |  |  |
| Questionnaire |  | X | X | X | X | X | X | X | X |
| CXR<sup>a</sup> |  | X |  |  |  |  | X | X |  |
| HIV test, CD4 count, viral load<sup>b</sup> |  | X |  |  |  |  |  |  |  |
| Clinical labs (e.g. CBC, HbA1c) |  | X |  |  |  |  |  |  |  |
| Sputum smear, culture, and GeneXpert Ultra<sup>c</sup> |  | X |  |  |  |  |  |  |  |
| Gastric aspirate<sup>d</sup> |  | X |  |  |  |  |  |  |  |
| Induced sputum<sup>e</sup> |  | X |  |  |  |  |  |  |  |
| IGRA and/or TST |  | X |  |  |  |  |  |  |  |
| Urine LAM |  | X |  |  |  |  |  |  |  |
| Whole blood<sup>f</sup> |  | X | X | X | X | X | X | X | X |
| Urine for storage |  | X | X | X | X | X | X | X | X |
| Spirometry<sup>g</sup> and/or Oscillometry<sup>h</sup> |  |  |  | X |  |  | X | X | X |
| 6MWT's<sup>i</sup> |  |  | X |  |  |  | X | X | X |
| Chest CT scan<sup>j</sup> |  |  |  |  |  |  |  |  | X |

<sup>a</sup> Chest x-ray (CXR) at baseline, unless done within 4 weeks prior to the Baseline Visit.
<sup>b</sup> HIV testing on participants not known to be positive to be performed per national guidelines; CD4 count and viral load will only be performed on participants who are HIV-positive
<sup>c</sup> For children whose diagnosis is made on the basis of a gastric aspirate (GA) or clinical criteria, subsequent GA inductions will not be required.
<sup>d</sup> Option for pediatric participants ≤10 years of age in whom there is high concern for TB, who do not have microbiologic confirmation of TB and are unable to produce spontaneous sputum
<sup>e</sup> Option for participants ≥3 years of age in whom there is high concern for TB, who do not have microbiologic confirmation of TB and are unable to produce spontaneous sputum.
<sup>f</sup> Whole blood processed to isolate plasma, serum, peripheral blood mononuclear cells (PBMCs)
<sup>g</sup> Children 5 years of age or older; baseline testing can be done at the Month 1 or Month 2 visit
<sup>h</sup> Children of any age; baseline testing can be done at the Month 1 or Month 2 visit
<sup>i</sup> 6-minute walk test in children 5 years of age or older
<sup>j</sup> Chest CT-scan will be performed at 18 months on children 5 years of age or older with persistent symptoms and/or abnormal CXR and/or abnormal spirometry

### Study Procedures at Baseline

Pre-enrollment screening will involve completing an eligibility form and informed consent. Study diagnostic procedures will be recorded in electronic case report forms. At baseline visits, participants will complete a symptom questionnaire. Medical history, physical examination, and demographic information will also be obtained. Anthropometric data will be collected to determine nutritional status. A chest X-ray (CXR) including lateral view will be done at the baseline visit. CXR will be read by both a study clinician and a radiologist using standardized reporting and scoring forms. Discrepant readings will be resolved by a pediatric pulmonologist. Tuberculin skin test (TST) and interferon-gamma release assay (IGRA) will be obtained at baseline visits. Respiratory specimen collection will include spontaneously expectorated sputum, or either induced sputum or gastric aspirate for children unable to expectorate. Each respiratory specimen will undergo GeneXpert Ultra testing, microscopy, and *Mycobacterium tuberculosis* (MTB) culture. HIV testing will be performed for children without documented HIV results, while children with previously confirmed HIV infection will have CD4 count and viral load testing. Whole blood and plasma will be collected for complete blood count analysis (CBC) and C-reactive protein (CRP). Urine will be collected for lipoarabinomannan (LAM) testing. Blood and urine specimens will be collected for future diagnostic studies.

### Study Procedures at Follow-up Visits

At each follow-up visit, participants will undergo a targeted symptom assessment and physical examination to evaluate clinical progress, and for those receiving treatment, response to anti-TB treatment. A repeat symptom questionnaire will be completed at every visit. Urine will be collected on all children at all follow-up visits. Children receiving INH for TB treatment or prevention will undergo urine INH metabolite testing. Remaining urine will be stored in aliquots for future analyses. Whole blood and plasma will be collected and stored at each visit for future analyses.

### Sample Collection

Sputum and gastric aspirate collection: sputum samples will be collected as spontaneously expectorated specimens when feasible. For children unable to expectorate, induced sputum will be collected using hypertonic saline nebulization. In very young children, or in cases where sputum cannot be obtained despite induction, early-morning gastric aspirates will be collected following overnight fasting using standard gastric lavage procedures. All sputum and gastric specimens will be transported to the laboratory immediately when collected at CRDR, or within 2 hours of collection when obtained at inpatient facilities, and processed for acid-fast bacilli microscopy, GeneXpert Ultra testing, and MTB culture.

Urine Collection: Urine specimens will be collected as clean-catch midstream samples when children are toilet-trained and able to cooperate. For infants and non-toilet-trained children, sterile adhesive collection bags will be applied to the perineal area after thorough cleansing with antiseptic wipes. For participants unable to provide samples at the study site, parents or guardians will receive training on proper collection techniques and will be instructed to return samples within 72-96 hours of collection. Bagged specimens will be monitored every 15-30 minutes and removed immediately upon voiding to minimize contamination risk. All urine samples will be processed within 2 hours of collection for LAM testing and stored appropriately for subsequent analyses.

Blood Draws and Maximum Volume Specifications: Venipuncture will be performed by trained phlebotomists using pediatric-appropriate techniques and equipment. Maximum blood volumes will be based on participant weight and age: children <2 years will have a maximum draw of 5 mL per session, children 3-17 years will have a maximum of 10 mL per session, with cumulative volumes monitored across all study visits. Blood collection will include whole blood for complete blood count analysis, plasma separation, serum isolation, and peripheral blood mononuclear cell (PBMC) isolation for future research applications. All blood draws will be scheduled to minimize patient discomfort and ensure adequate recovery time between collections.

### TB-specific Laboratory Procedures

All TB-specific laboratory procedures will be performed in accordance with CRDR standard operating procedures. GeneXpert MTB/RIF Ultra assay (Cepheid Inc., United States) will be performed at CRDR or the inpatient hospital facility where enrollment occurs. TB culture and speciation will be performed at a centralized laboratory facility. Liquid culture (BD BACTEC MGIT mycobacterial growth indicator tubes, Becton Dickinson Microbiology Systems, United States) will be used to detect growth. Samples will be inoculated into liquid culture per standard operating procedures depending on the type of specimen. If growth is detected, samples will undergo further testing with the BD MGIT TBc Identification Test (Becton Dickinson Microbiology Systems, United States), a qualitative chromatographic immunoassay for the MPT64 antigen to confirm presence of MTB.

### Evaluation of Novel Diagnostic Tests

Specimens obtained per this study protocol will allow for investigation of potential biomarkers of pediatric TB diagnosis and treatment response, including CRP, *M. tuberculosis* extracellular vesicles (Mtb-EVs), *M. tuberculosis* cell-free DNA, and other potential biomarkers. Studies are also planned to assess post-TB lung disease, treatment adherence, and CXR performance for pediatric TB with computer-aided detection software (CAD). **Table 3** provides an overview of these studies and associated specimens. The remainder of blood and urine samples will be kept in storage for future directions.

**Table 3.** Planned sub-studies.

| Study Name | Description and significance | Sample/<br>Investigation<br>type | References |
| --- | --- | --- | --- |
| Exosome | The utility of exosome-based Mtb biomarkers in pediatric TB has not been established. We will evaluate EV-associated antigens as a non-sputum diagnostic approach for children with presumptive TB. | Blood | 17 |
| CRISPR-TB | The role of CRISPR in pediatric TB diagnosis has not been established. We will evaluate its performance as an ultrasensitive, non-sputum assay. | Blood | 18 |
| CRP | The utility of CRP in pediatric TB management is not known. We will evaluate CRP as a diagnostic screening and treatment response tool | Blood | 19,20 |
| Adherence | Adherence data for TB treatment in children are limited. Adherence will be evaluated with a urine biomarker for isoniazid metabolites | Urine | 21 |
| PTLD | Risk factors for post-TB lung disease (PTLD) in children treated for TB are poorly understood. We will identify potential predictors of PTLD. | Spirometry,<br>Oscillometry,<br>CXR,<br>Computerized<br>Tomography<br>scans | 22–24 |
| CAD CXR | Computer aided detection (CAD) software is being used increasingly to screen for TB on chest x-ray. The accuracy of CAD software in diagnosing pediatric TB will be evaluated. | Chest x-ray | 25 |
| Lymphocyte-Monocyte | The monocyte-to-lymphocyte ratio (MLR) has been linked to active TB in adults and in HIV-infected pediatric cohorts, but its diagnostic utility in children with presumed TB more broadly has not been established. This sub-study evaluates whether blood-based MLR can distinguish confirmed TB from unconfirmed or unlikely TB and whether MLR changes with treatment. | Blood | 26,27 |

### Sample Storage

Several clinical samples will be stored at baseline visits and at follow-up visits including blood and urine stored at -80°C. Consent for sample storage will be obtained at enrollment as part of informed consent. Samples will be used in future evaluations of other novel TB diagnostics.

### Data Management

Data Collection and Storage: Data will be collected using Research Electronic Data Capture (REDCap) sponsored by the University of Washington (UW) Institute of Translational Health Sciences, with case report forms (CRFs) for data collection, storage, and management. REDCap will serve as the primary platform for data entry, storage, and management, providing password-protected access and secure data handling capabilities.

Quality Measures: We will implement quality assurance (QA) and quality control (QC) procedures throughout the study. This includes systematic review of research records for protocol compliance and Good Clinical Practice (GCP) standards. Site staff will conduct real-time monitoring and review all CRFs for completeness and accuracy.

Data Security: All data will be encrypted during transmission and storage. Access will be restricted to authorized study personnel including the principal investigator, co-investigators, and data managers. Regular database backups will be performed to ensure data integrity.

### Participant Confidentiality and Data Protection

We will assign each participant a unique study participant identification number (PTID) to be used on all study documents. No other identifying information will appear on data collection forms. Informed consent forms will be stored separately from study documents containing identification numbers. A password-protected file linking participant names to study identification numbers will be accessible only to senior staff. All study records will be maintained in locked files with restricted computer access.

### Clinical Case Definitions

The primary study endpoint is TB disease classification as per consensus criteria (Graham 2015)^11^ as described in **Table 4**, with modifications to include extrapulmonary TB. Participants who do not have appropriate length of follow-up to ascertain disease classification will be considered unclassifiable. Relevant data including clinical presentation, microbiologic testing results, response to treatment, and adjudicated baseline CXR interpretation will be presented to a two-clinician review panel with expertise in pediatric TB. Potential disagreements between reviewers will be resolved in further discussions including the study team. Study participants, treating clinicians, and study investigators will be blinded to the results of novel diagnostic assays during treatment and when assigning disease classification. We will create a STARD diagram for each investigational assay that includes the assay results and number of participants fulfilling each reference standard classification.

**Table 4.**
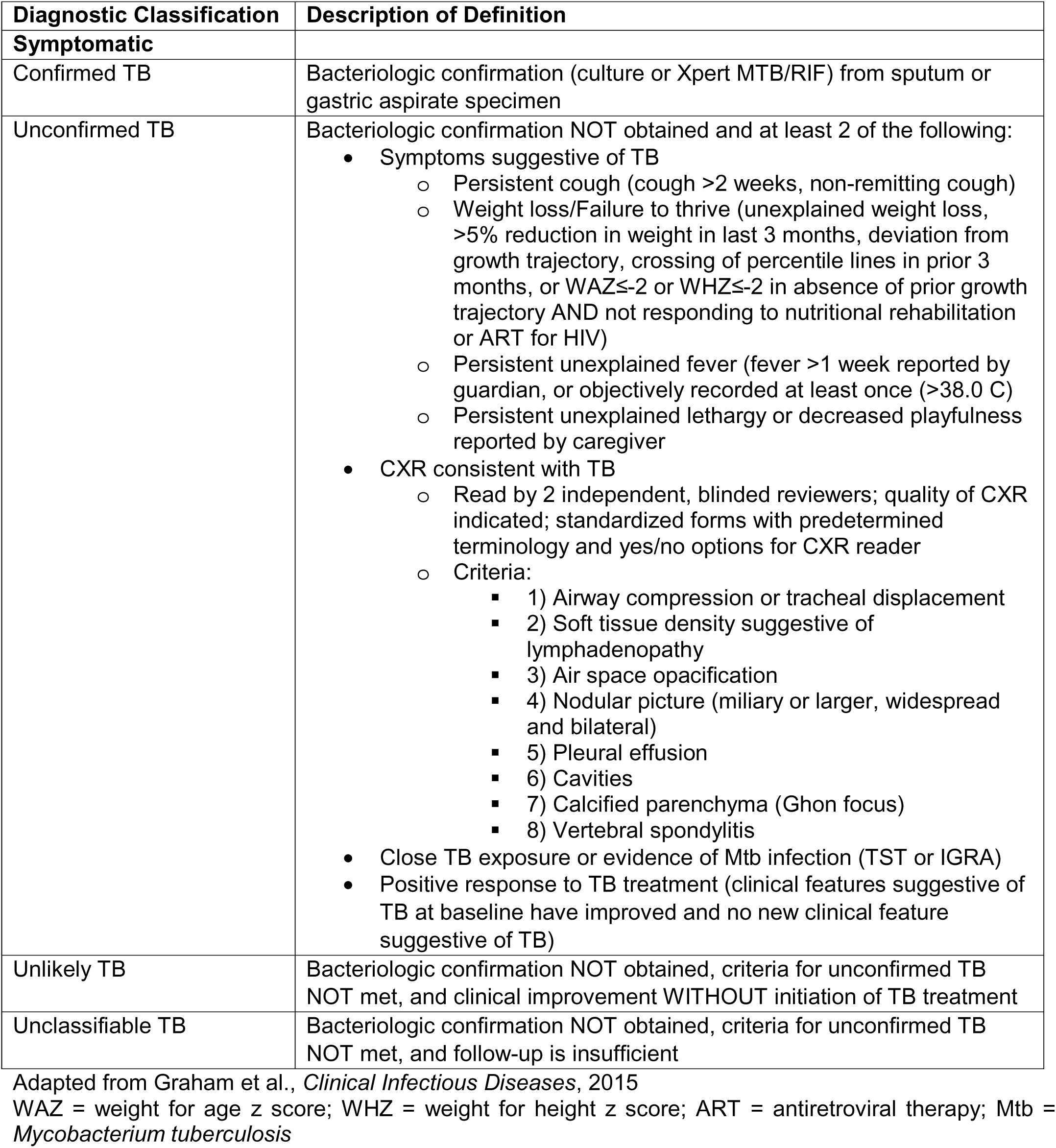
Case Definitions.

### Sample Size

This study will aim to include at least 400 participants presenting with TB which will allow for adequate estimations of the accuracy of novel assays. Assuming a 15% prevalence of confirmed TB in the cohort, an assay with 95% sensitivity would have a 95% CI of 86.1-99.0%, and 95.8-99.2% for 98% specificity.

### Planned Analyses

Primary analysis of novel diagnostics assessed in the cohort will include diagnostic performance as measured by sensitivity, specificity, positive and negative predictive values, and area under the receiver operating characteristic curve (AUC-ROC) using a composite reference standard of participants meeting criteria for confirmed TB or unconfirmed TB (**Table 4**). Sensitivity analyses will be performed to identify optimal sensitivity and specificity cut-offs for novel assays. Diagnostic performance and sensitivity analyses will be performed for the entire cohort and stratified by HIV status. To evaluate potential correlates associated with assay positivity, we will evaluate age, sex, TB disease severity, CXR results, nutritional status, HIV-related immune suppression, in multivariable modeling. We will also assess for the ability of novel diagnostics to detect early subclinical TB disease and their relationship with treatment response over time. Depending on the assay assessed, generalized linear models will be used to estimate association between treatment response and changes in biomarker over time. Treatment response will be defined as increase in weight and resolution of enrollment TB symptoms over the course of follow-up^12^. We will also use Cox proportional hazards regression to evaluate potential differences in mortality among participants with different biomarker characteristics at enrollment.

Because the diagnosis of pediatric TB relies on an imperfect reference standard, true prevalence is likely underestimated, and children with unconfirmed TB are often not included in assessments of new diagnostics. Bayesian latent class analysis is an increasingly important approach to estimate the accuracy of novel diagnostics in the setting of an imperfect reference standard and has been used in the evaluation of pediatric TB^13–15^. Using a Bayesian approach, we will estimate probability of “true” TB in the entire cohort, based on different assumptions of independence between test outcomes. We will also assess model variations with covariate adjustment for factors potentially influencing TB prevalence including age, sex, malnutrition, and HIV status. Using a Bayesian approach that includes the clinical information we anticipate will be available (TST/IGRA, CXR, treatment response), we will model conditional dependence between different tests. We will model TB prevalence and measures of diagnostic performance using informative priors of established assays (Xpert/culture and urine LAM) and likely non-informative priors of novel assays.

### Ethical Consideration

This study was approved by the University of Washington Institutional Review Board, the Kenya Medical Research Institute Scientific and Ethics Review Unit, and University of Nairobi-Kenyatta National Hospital Ethics and Research Committee. Study results will be published in peer-reviewed journals and disseminated to local TB treatment and screening programs.

## Discussion

The PDTBDx cohort study will aim to advance new methodologies and protocols for evaluating pediatric TB. Opportunities to include children and adolescents should occur early in the evaluation of novel diagnostic assays, as per recent pediatric TB consensus statement guidelines^16^. This study seeks to facilitate this goal by creating a large repository of specimens from children and adolescents with confirmed and unconfirmed TB over the course of treatment and observation.

Publication of this study protocol will allow for harmonization and consistency across pediatric TB diagnostic study cohorts, which is critical in the development of novel methods of diagnosing pediatric TB. Here, we describe the design, procedures, and definitions that will be used in the study. Participants undergo rigorous baseline testing and extended follow-up, consistent with internationally recognized guidelines for childhood TB classification. The extended 24-month follow-up strengthens diagnostic evaluation by allowing assessment across disease evolution, treatment, and recovery. The use of a consistent reference standard and disease classification schema further provides strength in analyzing new diagnostics that will be evaluated in the cohort.

Including both microbiologically confirmed and unconfirmed TB reflects real-world pediatric TB epidemiology, where bacteriologic confirmation is often limited. This enhances the applicability of diagnostic performance estimates to routine clinical care. Recruitment across inpatient, outpatient, and community-based facilities ensures representation across a full spectrum of disease severity and care-seeking pathways, enabling evaluation of diagnostic performance in settings reflective of routine practice.

The biospecimen repository, together with linked clinical and imaging data, will facilitate rapid evaluation of emerging biomarkers, molecular assays, and imaging-based diagnostic tools, thereby shortening the pathway between discovery and clinical validation. The longitudinal structure of the cohort further enables assessment of treatment response trajectories and post-TB lung health outcomes, areas that remain insufficiently characterized in children but are increasingly recognized as critical components of pediatric TB care and research. Although enrollment draws from multiple facilities across Nairobi County, study procedures are conducted at a single centralized site using standardized clinical and laboratory methods. This design provides strong internal validity prior to future evaluation of novel diagnostics in additional settings. Together, these features position PDTBDx as a valuable platform for advancing evidence-based tools to improve the accuracy, accessibility, and timeliness of pediatric TB diagnosis.

## Data Availability

All data produced in the present study are available upon reasonable request to the authors

## ACKNOWLEDGMENTS

The authors acknowledge the PDTBDx study staff for essential contributions to participant recruitment, clinical care, specimen collection and processing, data management, and imaging, health facility staff, Kenya Medical Research Institute (KEMRI), University of Washington (UW)-Kenya, and Kenyatta National Hospital Research and Programs operational staff. We also thank Dr. Tony Hu, Dr. Christopher J Lyon, Dr. Zhen Huang, and our Tulane University Center for Cellular and Molecular Diagnostics collaborators for study conceptualization and diagnostic assay design. Above all, our sincere thanks to the study participants.

## AUTHOR CONTRIBUTIONS

S.M.L. is the principal investigator and designed the study. V.N. is the site principal investigator. E.M-O. provided expertise on the clinical aspects of the cohort. J.M., J.G., and J.N.E. managed data collection and cleaning. B.T.M., S.M.L., J.N.E, and J.G. developed the tables and figures. B.T.M, S.M.L., J.N.E., and J.G. wrote the initial draft of the manuscript. All authors read and approved the manuscript.

## CONFLICTS OF INTEREST

The authors report no conflicts of interest.

## FUNDING

This work is supported by the NIH National Institute of Allergy and Infectious Diseases and the National Center for Advancing Translational Sciences (grant numbers NIH/NIAID R01AI162152 to S.M.L.; R01AI179714 to S.M.L; NIH T32 AI007044, B.T.M; NIH UL1TR000423 for REDCap; Tuberculosis & HIV Co-Infection Training Program in Kenya (TBHTP) (NIH Fogarty D43 TW011817), J.G.; University of Washington/Fred Hutch, Center for AIDS Research (CFAR) New Investigator Award (NIA) CFAR-NIA, J.G., (AI027757)

## Notes

### Competing Interest Statement

The authors have declared no competing interest.

### Author Declarations

This study was approved by the University of Washington Institutional Review Board, the Kenya Medical Research Institute Scientific and Ethics Review Unit, and University of Nairobi-Kenyatta National Hospital Ethics and Research Committee.

